# Seasonality and uncertainty in COVID-19 growth rates

**DOI:** 10.1101/2020.04.19.20071951

**Authors:** Cory Merow, Mark C. Urban

## Abstract

The virus causing COVID-19 has spread rapidly worldwide and threatens millions of lives. It remains unknown if summer weather will reduce its continued spread, thereby alleviating strains on hospitals and providing time for vaccine development. Early insights from laboratory studies of related coronaviruses predicted that COVID-19 would decline at higher temperatures, humidity, and ultraviolet light. Using current, fine-scaled weather data and global reports of infection we developed a model that explained 36% of variation in early growth rates before intervention, with 17% based on weather or demography and 19% based on country-specific effects. We found that ultraviolet light was most strongly associated with lower COVID-19 growth rates. Projections suggest that, in the absence of intervention, COVID-19 will decrease temporarily during summer, rebound by autumn, and peak next winter. However, uncertainty remains high and the probability of a weekly doubling rate remained >20% throughout the summer in the absence of control. Consequently, aggressive policy interventions will likely be needed in spite of seasonal trends.

## Main Text

Novel Coronavirus Disease 2019 (COVID-19) is causing widespread morbidity and mortality throughout the world *(1, 2)*. The SARS-CoV-2 virus responsible for this disease has infected over 2.2 million people when this article went into review *(3)*. Much of the world is implementing non-pharmaceutical interventions, including preventing large gatherings, voluntary or enforced social distancing, and contact tracing and quarantining, in order to prevent infections from overwhelming healthcare systems and exacerbating mortality rates *(2, 4)*. However, these interventions risk substantial economic damage and thus decisionmakers are currently developing plans for lifting them. Consequently, improved forecasts of COVID-19 risks are needed to inform decisions that weigh risks to both human health and economy *(2)*.

One of the greatest uncertainties for projecting future COVID-19 risk is how weather will affect its future transmission dynamics. SARS-Cov-2 might be particularly sensitive to weather because it survives longer outside the human body than other viruses *(5)*. Rising temperatures and humidity in the northern hemisphere summer could reduce SARS-CoV-2 transmission rates *(6-8)*, providing time for healthcare system recovery, drug and vaccine development, and a return to economic activity. Simultaneously, the southern hemisphere is entering winter, and we do not know if winter weather will increase COVID-19 risks, especially in developing countries with reduced healthcare capacity. Early analyses of COVID-19 cases predicted that high temperatures would reduce transmission during the summer *(9-11)*. These predictions have been widely reported in mainstream media and are informing decisions about relaxing control efforts soon. However, these analyses relied on the early stages of viral spread before the epidemic had reached warmer regions and thus potentially conflated weather with initial emergence and global transport.

We estimate how weather affects COVID-19 growth rate using data through April 13^th^, 2020 by applying methods that improve model predictive accuracy, incorporate uncertainty, and reduce biases. Based on emerging evidence, we developed several *a priori* predictions about how weather, either directly or indirectly via modified human behaviors (e.g., aggregating indoors), affects COVID-19 growth rate. Preliminary research on SARS-Cov-2 *(9, 10, 12)* and related viruses *(8, 13)* predicted that COVID-19 growth would peak at low or intermediate temperatures. However, other coronaviruses demonstrate weak temperature dependence, instead depending on social or travel dynamics *(7)*. High humidity also might decrease viral survival, limit transmission of expelled viral particles, or decrease host resistance *(13-15)*. Ultraviolet light effectively kills viruses such as SARS-Cov-1 *(16)*, and thus sunny days might decrease outdoor transmission or promote immune resistance via vitamin D production *(17)*. We also evaluate demographic variables, assuming greater transmission in denser and older (>60) populations.

We modeled maximum growth rate of COVID-19 cases to estimate contributions from underlying climate and population dependencies without healthcare interventions (e.g., social distancing). Hence, we restrict analyses to the early growth phase before interventions reduced transmission, but after community transmission began, when the vast majority of the population was still susceptible to this novel virus. We estimated the average maximum growth rate *(λ)* as the exponential increase of cases (ln(N_t_) – ln(N_0_)/t, where N_*t*_ = cases at time, *t*, and N_0_ = initial cases) for the three worst weeks in each political unit (country or state/province depending on available data *(3)*), where *t* = 7 days (see Supplementary materials for additional periods). Testing and reporting of COVID-19 likely vary across political units. However, estimated growth rates should remain robust to these biases assuming detection probabilities remain constant during the short, one-week estimation period. We restricted analyses to locations with >40 cases to eliminate periods before local community transmission. Applying these criteria, we used data from 128 countries and 98 states or provinces.

We applied Bayesian Markov Chain Monte Carlo methods with uninformative priors to estimate parameters. We obtained daily infection data from *(3)* and 3-hour weather data from the ERA5 reanalysis for the 14 days preceding case counts consistent with the 1-14 day infective period *(18)*. We used fine-scaled weather data rather than long-term climatic monthly means to model observed weather-outbreak dynamics. Weather data was weighted by population size in each 0.25° grid cell within each political unit to capture the weather most closely associated with outbreaks in population centers. We used leave-one-out cross-validation to choose the best models, which ranks model on predictive accuracy on excluded data. We included a random country effect to account for differences in national control response times, health care capacity, testing rates, and other characteristics intrinsic to country of origin.

The best model for predicting maximum COVID-19 growth rate predicted 36% of the variation in COVID-19 growth rates (Fig. 1), and 17% excluding country effects. This model included maximum daily ultraviolet light, mean daily temperature, proportion elderly, and mean daily relative humidity (Fig. 2A). Competing models reflected the same qualitative results and similar parameter estimates (see Supplementary materials). Ultraviolet (UV) light had the strongest and most significant effect of tested meteorological variables on COVID-19 growth *(β*_*UV*_ = −0.44, 95% credible intervals (Cis): −0.53, −0.36). Contrary to predictions, temperature positively affected COVID-19 growth rate *(β*_*temp*_ = 0.23, 95% CIs: 0.15, 0.32), although this is conditional on accounting for UV in the model (note that temperature and UV are moderately correlated (r=0.75) in our data set and extensive testing was done to ensure coefficients estimates were not an artifact of this correlation; see Supplementary Materials). As expected, relative humidity decreased growth rates, although not significantly, either by reducing the virus’ survival outside humans or reducing airborne transmission *(β*_*humid*_ = −0.05, 95% CIs: −0.11, 0.00). Absolute humidity was strongly correlated with temperature *(r* = 0.88) and thus could be exchanged with temperature with little difference in model performance. Contrary to predictions, the proportion of elderly decreased COVID-19 growth rate *(β*_*popsize*_ = −0.07, 95% CIs: −0.14, - 0.00), most likely due to outbreaks in developed countries with older populations. Population density was not selected in any top model. The model was characterized by equally strong random effects associated with country of origin (Fig. 2B). For instance, Turkey, Brazil, Iran, the U.S., and Spain had the highest growth rates independent of modeled factors, whereas China, Iceland, Burkina Faso, Sweden, and Cambodia had the lowest. The strong negative effect associated with China indicates the effect of early interventions and is accounted for in our model. Notably, while intervention will substantially influence the absolute values of growth (i.e., the intercept in our model), our predictions can still be interpreted to represent relative differences in risk throughout the year.

**Figure 1.**
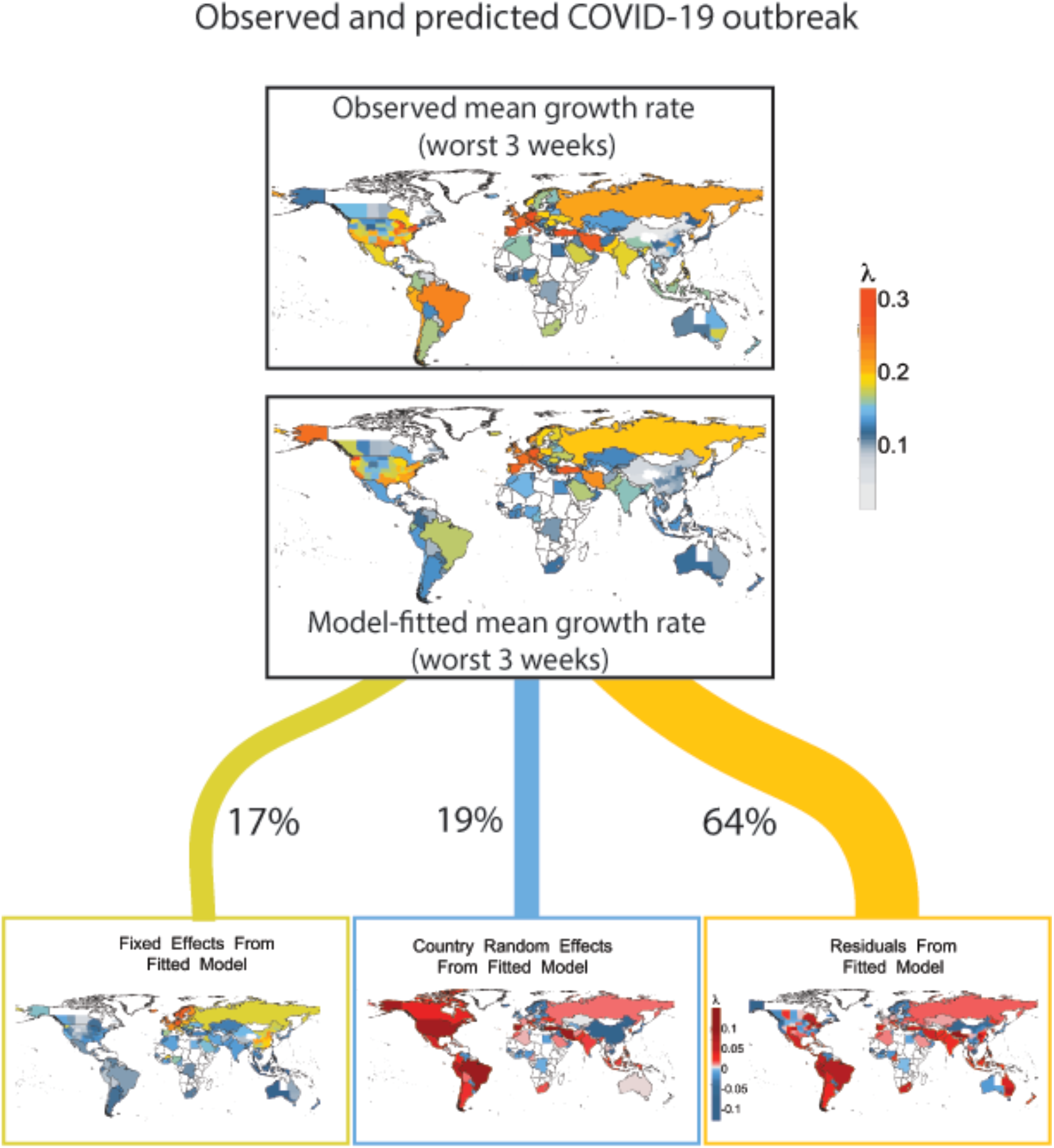
Observed and predicted maximum growth rates for COVID-19 along with graphical partitioning of model components including (A) weather and demography, (B) country effects, and (C) residual variation. Country effects are shown as the difference in growth rate between the country and the global mean. Only 17% of variation is explained by seasonality, while 19% of variation arises from country specific factors which may include quarantine policies, healthcare, or reporting practices.

**Figure 2.**
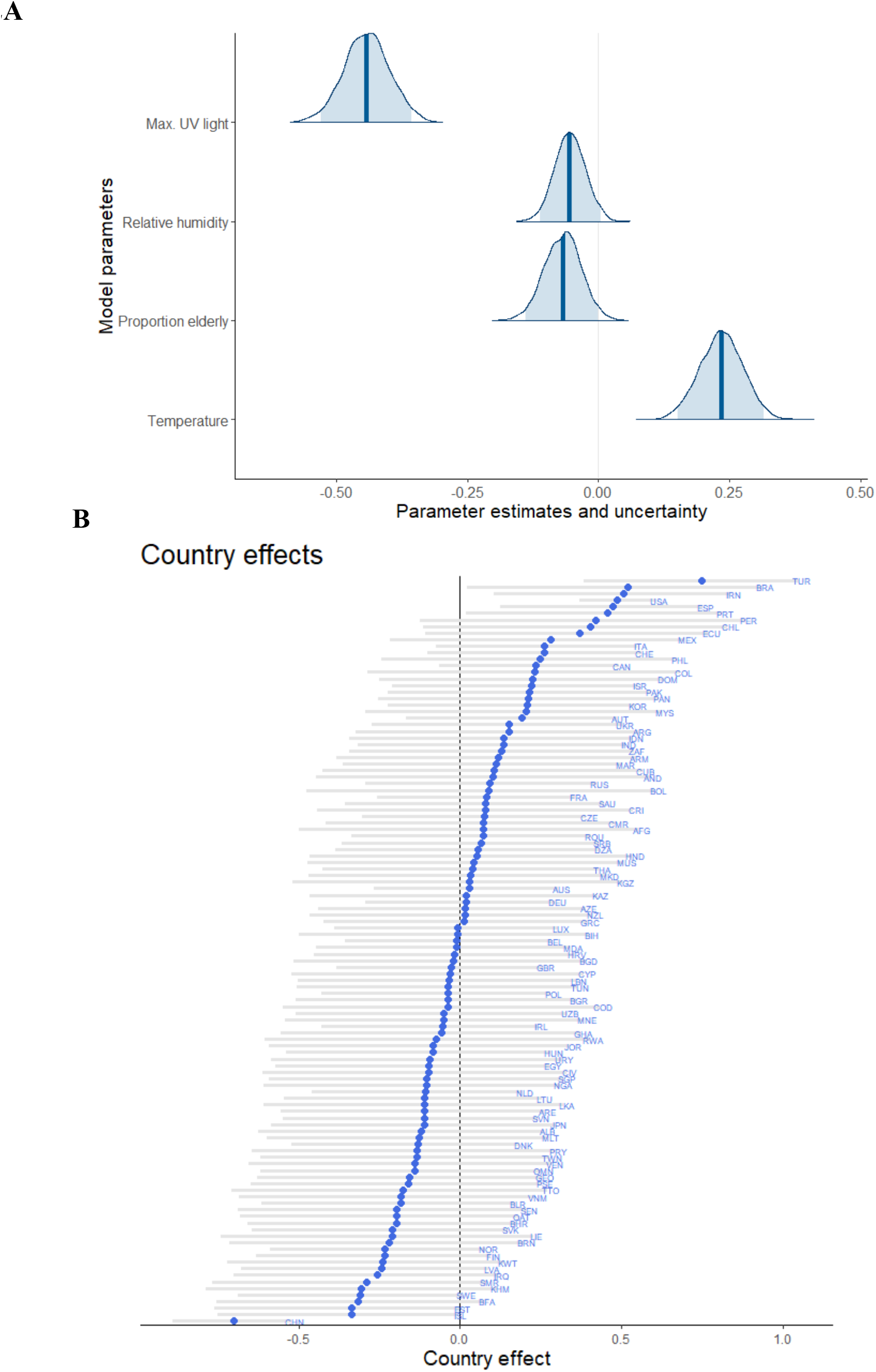
Median standardized estimates for weather and demographic factors (A) and for country effects (B) for best predictive model with 95% credible intervals (light blue). Country codes in B follow GADM ISO3 conventions.

We next explored why earlier studies might have predicted a negative association between temperature and COVID-19. Alone, temperature has a weak, negative effect on COVID-19 growth rate in our model, which becomes positive after adding other factors, and in particular, UV. Even with other parameters, temperature negatively affects COVID-19 early in the pandemic (Fig. 3, top). Significant positive temperature dependence emerges by late February following transmission to warmer, high-UV regions of climate space, like Africa *(19)* (see Fig. 3 bottom for filling of climate space). Notably our analysis does not specifically attempt to reproduce previous studies, so differences are expected depending on the details of decisions in other studies. This finding urges caution in estimating climatic niches of new, pandemic pathogens before they reach an equilibrium distribution with climate. Initial climate associations with viral outbreaks will first correlate with the narrower range of climatic variation found at the emergence site and then in global transportation hubs, rather than reflecting ultimate biological limits on growth and survival. We recognize that future data could alter our predictions further, especially as COVID-19 becomes endemic *(15)*. However, less variable model predictions and exposure to the most common global climates by April (Fig. 3) suggest that model predictions might have stabilized at least for now.

**Figure 3.**
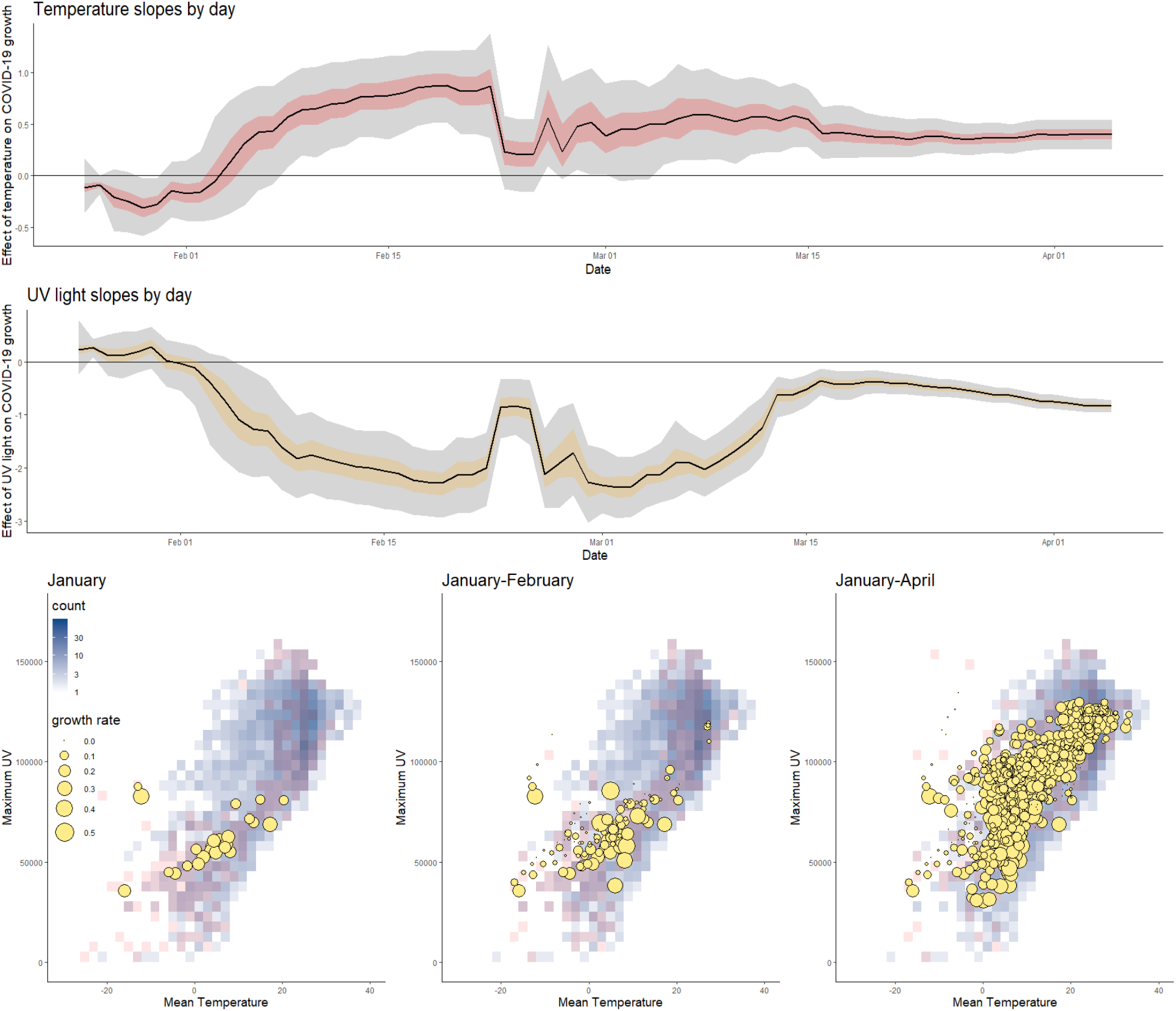
The effect of temperature and UV on COVID-19 growth rate as the pandemic spreads to new climates. Top, early COVID-19 outbreaks (indicated by growth rate proportional to blue symbol area) occurred in a subset of potential temperatures (°C) and ultraviolet light (Joules/m^2^) levels possible in a year (background gray-blue gradient) and for that specific time period (red overlay) based on counts of 5-year averages of climate variables. Bottom, Model coefficients and uncertainty through time demonstrates dynamic shifts and stabilization of parameter estimates (50% and 95% credible intervals indicated by colored and gray fills) and illustrates how earlier studies may have detected a negative temperature dependence. Before Feb 24 patterns were dominated by data from China, until large jumps in cases in Iran, Italy, and Japan appear in the data set, providing novel climate space to inform the model and leading to an abrupt change in model coefficients.

Using our model, we predicted potential COVID-19 growth rates in the upcoming months relative to a weekly doubling rate *(λ*=0.1; Fig. 4). Based mostly due to variation in UV and temperature, our model predicts that COVID-19 risk will decline across the northern hemisphere this summer, remain active in the tropics, and increase in the southern hemisphere as days shorten and UV declines (Fig. 4, left and right panels). However, given high uncertainty, a non-negligible risk exists throughout the world for potential outbreaks in summer similar to that observed at the outset of the pandemic (Fig. 4, middle panel, dark blue = 30% probability of *λ*>0.1). By September, declining daylength steadily increases predicted risks of COVID-19 outbreaks in the northern hemisphere until a peak in December-January, while risks decline in the southern hemisphere. Although this model represents our best current estimate, a range of outcomes still remain possible (Fig. 4 middle). Furthermore, these predictions of potential growth need not be realized if appropriate interventions are enacted or a vaccine is developed. The overall conclusion is that although COVID-19 might decrease temporarily during summer, there is still a moderate probability that it is weakly affected by summer weather, and that it could return in autumn and pose increasing risks by winter.

**Figure 4:**
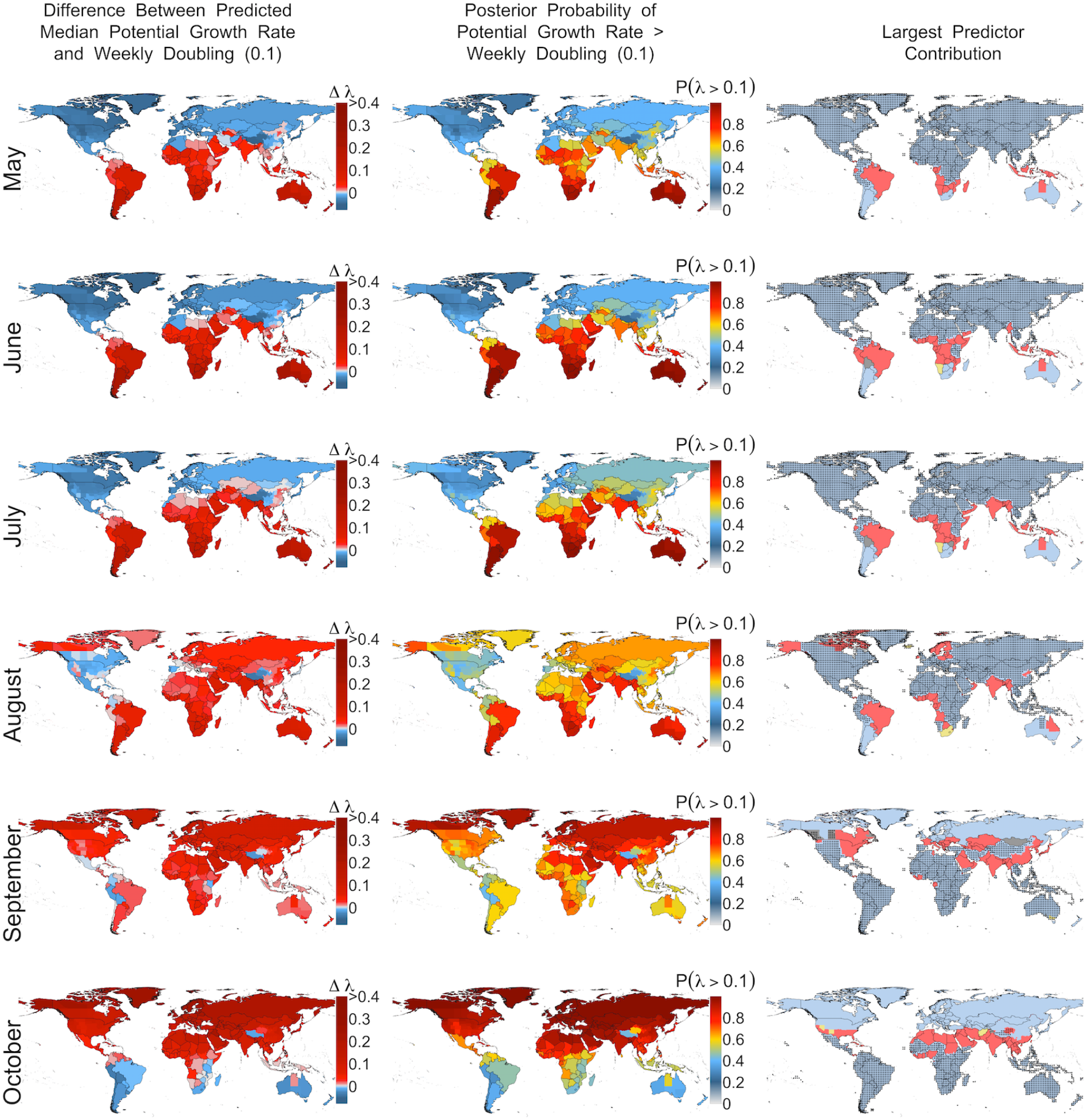
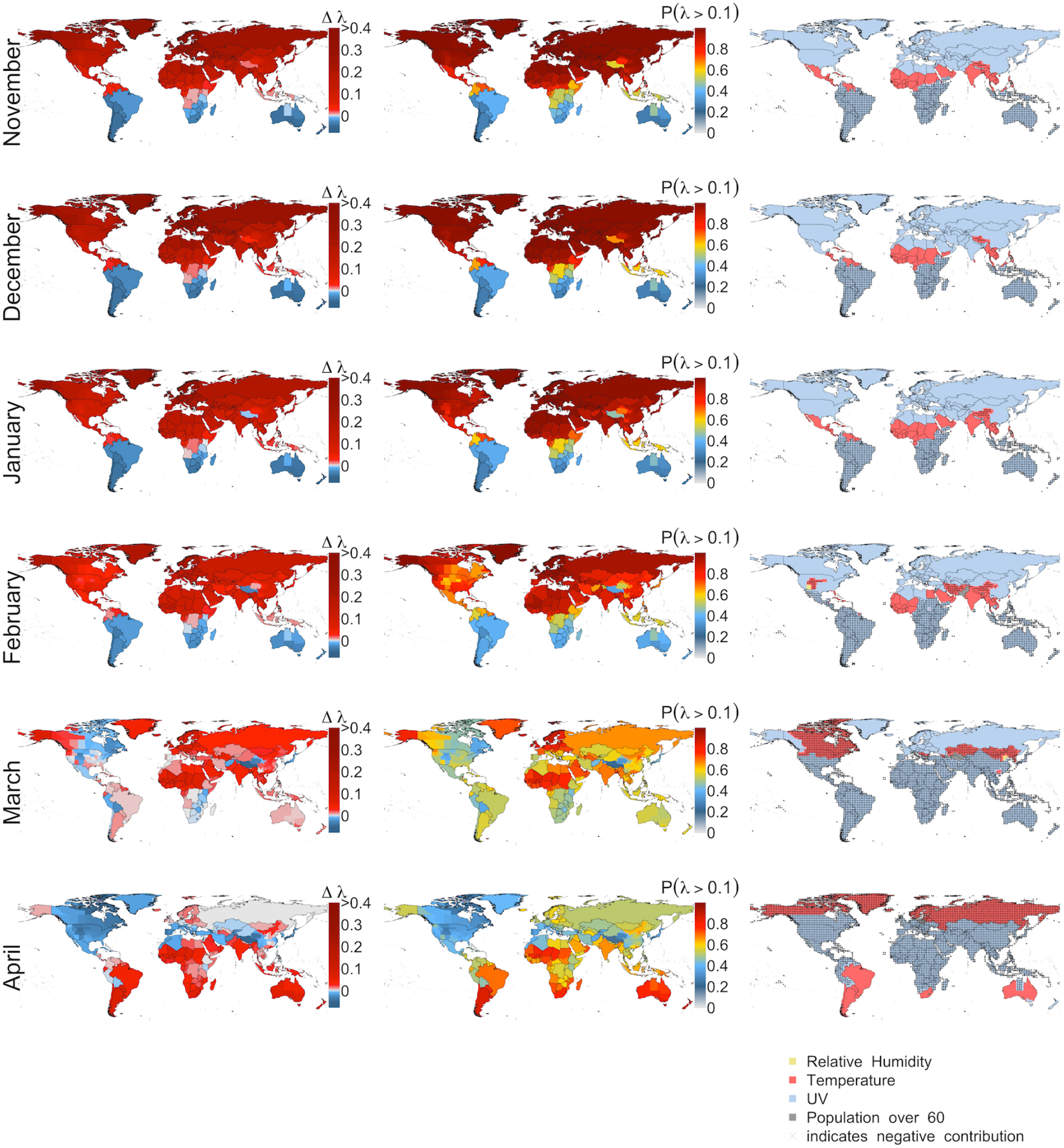
Predicted potential growth rates of COVID-19 by month using best model. Leftmost column, Potential growth rate relative to weekly doubling time *(λ* = 0.1). Red indicates faster than a weekly doubling rate and blue indicates slower rates. Central column indicates the posterior probability of growth rates exceeding a weekly doubling rate. Rightmost panel indicates which predictor contributes most (based on predictor*coefficient) to COVID-19 growth rate *(λ)* in each 0.25° cell, with stippling indicating negative contributions.

Our predictions were robust to the manifold decisions made regarding data and model structure. We explored the consequences of using different parameter comparisons, the effects of shorter (7-day) time windows for aggregating weather data, different cut-offs for minimum number of cases, varying number of weeks analyzed per political unit, whether we analyze the first or worst weeks following the infection threshold, and if we included weather maxima and minima instead of means and found no qualitative changes to results, with the exception that maximum daily UV during at 14-day interval substantially outperformed the mean (Supplementary materials). We also explored the effect of excluding data from China, which lacked data prior to control measures in many cases, and found similar results.

Understanding the true contributions of weather to human pathogens requires combining insights from observational analyses like this one and manipulative experiments that isolate factors under controlled conditions *(5, 12)*. Other causal factors correlated with weather variables in our model could have contributed to our findings, including weather-associated human behaviors (e.g., seasonal aggregations for education or religion). Despite initial suggestions that seasonality would strongly control COVID-19, weather only explains 17% of the variation in COVID-19 growth rates. Undescribed factors at the level of political units were just as important as weather (19% of variation), and much of the variation (64%) remains unexplained. Future studies should embed these meteorological insights into epidemiological models that include human demography, movement, sociocultural behaviors, healthcare capacity, and political interventions [e.g., *(2, 4, 15)*].

We demonstrate that COVID-19 growth rate increases with reduced ultraviolet light, higher temperatures, and lower relative humidity. We predict that COVID-19 will oscillate between the northern and southern hemisphere, based largely on seasonal variation in UV radiation and temperature without continuing interventions like social distancing. Despite a possible, but highly uncertain, temporary summer reprieve in the north, COVID-19 is more likely to return by autumn and threaten further outbreaks. The north should take this time to build resilience against future outbreaks, while assisting countries in the tropics and southern hemisphere. Uncertainty remains high, however, so we urge caution when making decisions such as removing societal interventions before more permanent pharmaceutical solutions can be implemented. Overcoming this pandemic will take extensive global collaborative scientific efforts to unravel its biology as well as the continuing resolve of people worldwide adhering to social restrictions.

## Data Availability

All data come from public data sources.

## Acknowledgments

NSF awards HDR-1394790 supported CM and DEB-1555876 supported MCU. We thank the Center for Systems Science and Engineering (CSSE) at Johns Hopkins University for providing COVID-19 data.

## Methods

### Overview

We examined the weekly rate of increase in the number of COVID-19 infections as a function of weather, while controlling for human population structure, in order to determine the effects of the abiotic environment on the growth rate of infections (). Our selection of weather variables and the time frame within which we measured variation was based on the limited, but rapidly expanding, experimental and observational research on the survival and transmission of SARS-CoV-2 virus and human resistance to the resultant COVID-19 disease *(1–5)*. We performed model selection to optimize model prediction of cross-validated data and performed comprehensive sensitivity analysis with respect to both data preparation and modeling decisions and found no qualitative differences between the findings represented in our best model and other models using different, but reasonable, decisions.

### Infection Data

Daily infection data were obtained from the Johns Hopkins Center for Systems Science and Engineering *(6)*, which documents country level aggregations of infected individuals, except in Australia, Canada, China, and USA, where state-level data are available. From these daily data, we calculated weekly growth rate () assuming an exponential model for the growth of the number of infected individuals, which fit well to COVID-19 dynamics during the early stages of spread. The starting point for one-week intervals were polity specific (either country or state level depending on the resolution of available data) and calculated beginning on the first day (denoted t0) that the number of infected individuals exceeded 40 (and 20 and 60; see sensitivity analysis below). This minimum was necessary to eliminate the early dynamics of COVID-19 in locations due primarily to transport from other regions rather than local, community transmission. This moving window approach allowed us to capture local differences in onset date of transmission without imposing any artificial cutoffs (e.g., based on calendar week). By summarizing the data in this way, we had 541 observations of distributed over 203 political units.

To capture periods when the spread rate was most severe, we chose to focus on the worst three (also two, four; see sensitivity analysis) weeks in each political unit based on the magnitude of lambda, for our model. We were primarily concerned about high rates of spread, and their possible drivers, so this decision controls for differences among polities in the onset of severe spread and differences in the timing of control measures that may reduce growth. Hence, a focus on maximum growth rates is the best, unbiased estimate of COVID-19 growth in the absence of control measures, and most likely to be influenced by weather. In sensitivity analyses, we also considered using the first 2,3, or 4 weeks following t0, and found similar, but more noisy results, owing to the likely variation among countries in the early rates of spread (e.g., in Thailand, growth was initially low before increasing rapidly).

### Weather data

Weather data was aggregated from 3-hourly data downloaded from the ERA5 model by ECWMF *(7)* and averaged at 14 day intervals preceding the time period in which calculated for each polity. A 14 day interval captures the known infective period of SARS-CoV-2, where infections are known to occur from period of one to 14 days *(8)*. Hence, we use the actual observed weather during the period of viral transmission. This decision contrasts with previous studies that used average monthly climate calculated over the interval 1970-2000 provided by Worldclim *(9)*. Notably, the biweekly averages we calculated are, on average, expected to reflect higher temperatures due to climate change in the last 50 years compared to historic long-term averages. Further, our biweekly estimates better reflect that actual conditions when infections occurred, and thus are expected to better predict transmission if indeed they influence it.

Based on existing insights about SARS-CoV-2 and the onset of COVID-19, we considered the following weather variables: temperature 2m above land surface, relative humidity, absolute humidity, and total incoming UV radiation at the land surface. To align the weather data with infection data for a given political unit, we determined the first day (t0) when more than 40 individuals were reported (also 20 and 60 infections; see below). We calculated the mean values of the weather variables over the 14-day window preceding t0. For example, t0 for Connecticut, USA was March 16 (when 41 records had accumulated), so the weather variables were averaged over the 14-day window preceding March 10. This reflects the assumption that detected infections between March 10-16 primarily occurred between February 24-March 9. Although imperfect, the temporal autocorrelation of weather suggests that this is reasonable (e.g., even if an infection occurred on March 2, there is typically high correlation on weather, then, and March 3-9).

Finally, note that we also explored the use of minimum and maximum values of weather variables to account for the possibility that transmission was more likely driven by extreme weather rather than average weather. We also considered using weekly rather than biweekly intervals to reflect the possibility of shorter incubation periods. Outcomes were robust to these decisions.

Previous studies have noted that the coarse spatial grain of infection data (country or state level) makes it difficult to interpret weather variables in the context of such large spatial units *(10)*. To address this, we calculated weather averages over the quarter degree grid cells in a polity, weighted by the population size in each cell. This resulted in weather covariates that better reflect where most humans are and hence where infections occurred. Also, early maximum transmission rates were usually located in large cities, and thus weights weather variation in line with this bias.

### Population data

We obtained human population data from Worldpop.org focusing on total human population (density) and proportion of the population over age 60. Population density was hypothesized to control for the number of interactions individuals in a location were likely to experience whereas the proportion of people over 60 in a polity was hypothesized to control for reporting rate, given that older people are more adversely affected by the disease and thus more likely to be tested.

Data were obtained at 1km resolution and summed to the quarter degree grid imposed by the weather data. Polity information was obtained based on global standards (GADM.com). Each quarter degree grid cell was assigned to a polity and cells were averaged over the polity.

### Models

We focus on the growth rate of COVID-19 cases,, rather than estimating a climate niche for the virus based on its presence or absence or total number of cases, as explored in preliminary studies *(11)*. to avoid issues with disequilibrium in the virus’ distribution. We focused on estimating the rate of increase () of infected individuals, rather than directly modeling the number of infected individuals, in order minimize the influence of different reporting biases in different polities. We calculated as= (ln(N(t)) -ln(N(t0))/t where t was taken to be 7 days and t0 defined the start date for counting infections. This formulation is independent of reporting bias under the assumption that the reporting bias is constant over the 7-day interval. To see this, consider that the true number of infected individuals N* is related to N via the proportion of cases reported, p, such that N=pN*. Substituting this expression for N into the expression for, it is apparent that p cancels out. Hence so long as p is approximately constant across a 7-day interval, it does not affect the estimate of growth rate.

We modeled with a hierarchical Bayesian Gaussian regression with a log link on the weekly transmission rate. The full model included mean 14 day lagged temperature, mean 14 day lagged relative humidity, mean 14 day lagged absolute humidity, mean 14 day lagged UV, human population density and proportion of the population over 60. We used linear terms for all variables but also considered a quadratic term for temperature based on suggestions of modality in previous studies *(11–13)*. Based on sensitivity analyses discussed below, we found that maximum daily UV was a considerably better predictor than the mean (delta LOOIC = 4.x) so we used the maximum in our best model. Country-level random effects were used to capture differences in policies, health care or other locally specific behaviors. We also explored state/province-level random effects (where applicable), but country-level effects performed considerably better in all models explored based on model selection criteria.

### Model selection

We were interested in developing models with high predictive ability. Thus, we performed model selection using leave-one-out (LOO) cross validation. This technique iteratively uses all data except for the ith data point to develop a model, then it predicts the left-out point, and uses the divergence between model prediction and observation to rate model performance. The sum of these divergences across all N data points is then converted into a standard measure of overall model performance called the Leave-One-Out Information Criterion (LOOIC), where lower numbers indicate models that better predict left-out data *(14)*. This model selection method has been found to excel over alternative Bayesian methods such as Deviance Information Criterion, and is especially appropriate when the objective is prediction *(14)*.

Model selection was performed by starting with the full model and using forward and backward stepwise selection. The full model regressed the growth rate over a one-week window against linear terms for mean temperature, mean UV, mean relative humidity, mean absolute humidity population density, and proportion of the population over 60. We included a quadratic term for temperature based on earlier studies suggesting a decline in of growth rate with temperature. We also included an interaction term between temperature and UV to account for their correlation. All these variables were calculated in the 7-day windows preceding the interval used to calculate growth rate. During stepwise selection, we note that there were no cases of parameters trading off with one another and that coefficients for each predictor always retained the same sign and approximate magnitude regardless of which other predictors were in the model. The only exception to this was when UV was excluded from a model that included temperature; the temperature effect dropped from positive to near zero. Hence it is important to interpret the positive effect of temperature in our best model as accounting for the effect of temperature only after UV has been included in the models.

Once we found the best suite of predictors (excluding the quadratic temperature term, the UV-temperature interaction, absolute humidity, and population density), we explored whether using the maximum or minimum daily values of each weather variable, and 7 versus 14 day lagged intervals, improved LOOIC. The only case where we found significant model improvement compared to the biweekly means was for maximum UV over both 7- and 14-day intervals. Since the 14-day interval improved model performance most (based on LOOIC), we chose that as the summary statistic for UV. Notably for all other weather variables there was negligible difference in LOOIC when we used weekly versus biweekly means and hence we used biweekly values for all variables for simplicity.

### Sensitivity analysis

Sensitivity analysis for a variety of model decisions was conducted to determine whether our key finding - the relation between COVID-19 growth rate and temperature, UV, and relative humidity - was affected by any of our decisions. In all cases that follow, the median of the temperature coefficient was positive, with a 95% credible interval sometimes overlapping zero and sometimes not, depending on the model. In all cases, the median and 95% credible interval for UV was negative. In all cases, the 95% intervals for relative humidity and population density always overlapped zero but the medians were always negative and positive, respectively. The quadratic temperature term never improved the model, indicating that there was no support for a unimodal response to temperature.

Sensitivity to a number of data preparation steps was assessed. During data preparation, we considered the 2, 3, and 4 worst weeks (highest lambda) following t0, as well as the first 2,3, and 4 weeks following t0. We chose different cutoffs (20,40,60) for numbers infected to account for the difficulty in determining the time when spread became local, rather than imported. Due to the strong control measures in place in China by the time our data set begins (January 22, 2020), we also compared our best model with and without data from China and found no qualitative change in outcomes.

### Coefficients over time

To explore how our inference about different weather factors may have changed over time, as the virus approaches a geographic and environmental equilibrium (which it may still not be at), we fit a model each day since February 1, 2020, accumulating infection data up until the most recent date of analysis. This analysis can illustrate (1) how earlier studies may have inferred a negative dependence of growth on temperature, (2) the uncertainty inherent in earlier estimates of temperature dependence, (3) the disequilibrium between COVID-19 and the environment early in its spread, and (4) the smaller credible intervals, and hence increasing confidence in our model based on more recent data. Note that the model used to illustrate this pattern (1) used the first (rather than the worst) 3 weeks following t0 to accumulate data as early as possible and thus reflect decisions made in earlier studies, and (2) used polity (rather than country) effects because the data in early February was predominantly from China and thus country effects could not be fit. Although early data gaps meant that we could not precisely replicate previous analyses with this exercise, we obtained similar outcomes using this model for the present analysis (again indicating model robustness). As well, this exercise demonstrated how conclusions from earlier studies may have arisen, even with our more refined model, but based on a longer time series.

### Projections

Future predictions of the *potential* growth rate in May-September was made by projecting our highest performing model according to LOOIC. Importantly, we reinforce that our predictions pertain to the possible growth rate in the absence of social distancing or other control measures because it is based on a model fit with infections that occurred primarily before precautionary policies were implemented. Note that even if a policy were implemented on, for example, March 14, we expect that infections reported in the next two weeks were initiated before the policy began. Hence, we predict the underlying contribution of weather to future COVID-19 growth. Importantly, these predictions reflect what would happen if other control measures are relaxed and the natural dynamics of infection can begin again in a population with little resistance. Currently governments are deciding when and how to relax control measures, often under the assumption that weather will lessen the potential for spread in the upcoming months. Thus, whereas we do not presume to predict the actual future growth rate of COVID-19, we do hope to capture the potential maximum growth rate in order to inform the relative risks of alternative control strategies.

To make future projections, we obtained monthly mean temperature and relative humidity weather data from 2015-2019 from the same data source as above, under the assumption that these recent years are representative of what to expect in the coming months. Notably hotter or cloudier (lower UV) days in the coming months would suggest higher growth rates than we predict. UV data was not available in a monthly aggregation, so we obtained the 3-hourly data and aggregated it to monthly values. Human population was assumed to remain constant. We projected the models without random effects (or equivalently at the mean value of 0) as we were reluctant to assume that country-level policies, reporting, or health care potential will remain the same in the future. We expect that different country-level effects will dominate in the future, but predicting these offsets is beyond the scope of this study.

### Caveats

As with any predictive study, we seek to use the best available data and understanding of mechanisms to develop possible projections that make clear underlying decisions and uncertainty. Ultimately, such predictions must be treated with appropriate caution given the limited understanding of SARS-CoV-2 virus, human resistance, and its transmission dynamics at this time. Thus, while we seek to inform decisions, those decisions must also recognize the inherent uncertainty in any predictive model, but especially in the context of limited information. Future data will ultimately be the arbiter of these predictions, and thus good predictive modeling will require repeated bouts of model validation, revision, and re-projection as we learn more about this virus.

In particular, we await mechanistic information on viral physiology and human resistance to move beyond the correlative approach taken here by necessity. Mechanistic models apply insights about an organism’s intrinsic biology using parameters often collected from careful experimental manipulations. However, in the absence of this information, correlative models can predict near-term dynamics with accuracy *(15)*. Bayesian approaches like ours can integrate both mechanistic and correlative knowledge as these pieces of information become available.

One thing that we do not account for in our model is human behavior and control measures. By modeling maximum growth rate and using a threshold number of cases, we restrict our analyses to the period during which the disease expanded quickly, following the beginning of community transmission but before major control measures were implemented. For instance, most countries began implementing national control measures in mid-March, which would influence infections recorded into early April, based on a 14-day window for symptoms to emerge. Hence, we chose to limit our data set to records before April 7. However, we note that following early April, growth rates are expected to be much lower due to control measures, and these will continue to be important to reduce growth rates below the potential values we predict here which do not account for control.

We used available insights about SARS-Cov-2, related viruses, and observations of COVID-19 dynamics to select a list of factors that likely influence it. Although we purposefully limited these variables to reflect our best knowledge and to avoid overfitting, certainly other climate and epidemiological factors are likely missing from the model. Future studies should consider embedding these climate insights into epidemiological models that include human demography, immunity, movement, behaviors, medical capacity, and control efforts *(4)*.

## Supplementary Materials

**Figure S1.**
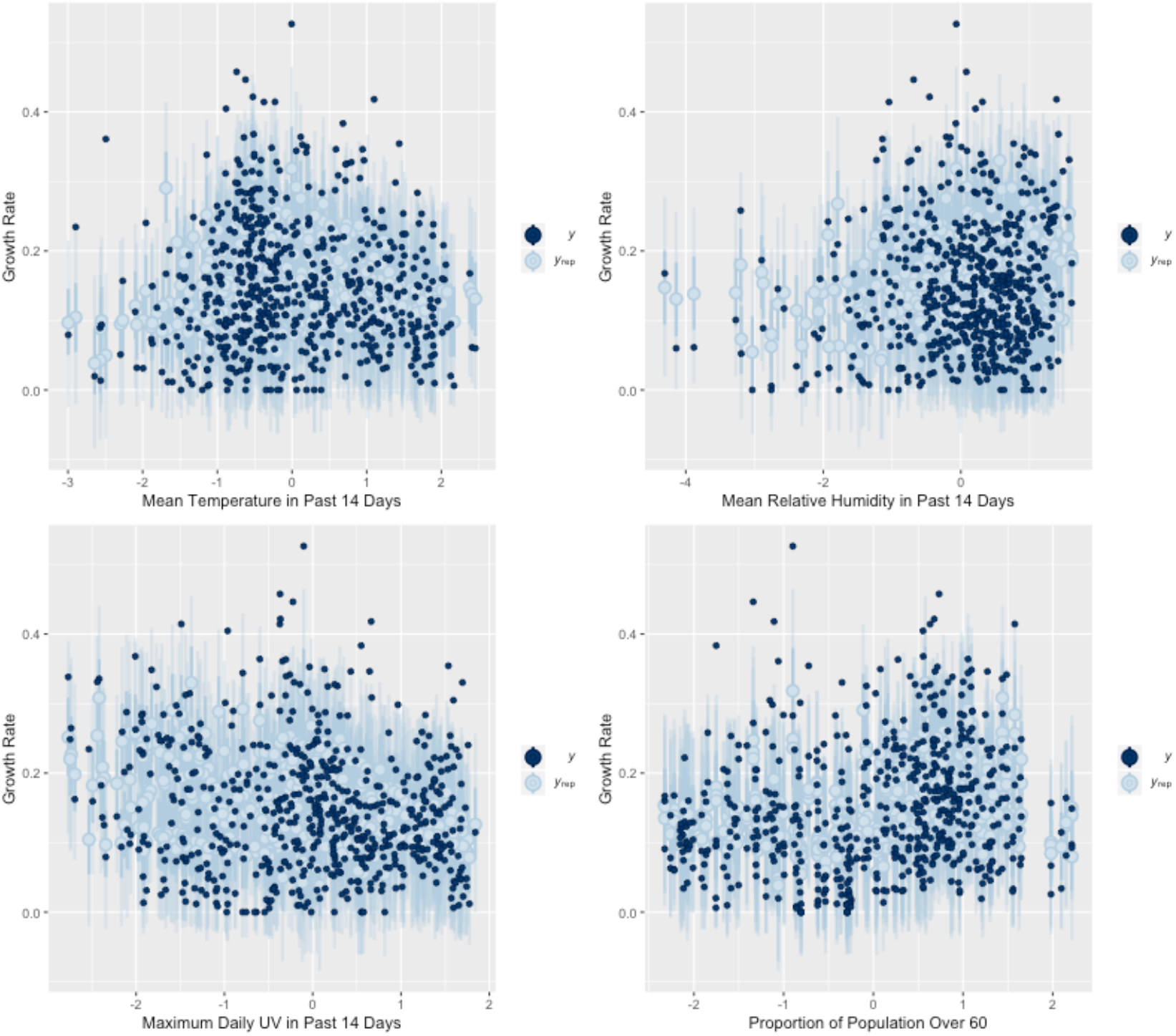
Posterior predicted probabilities of growth rate refelect weak trends with environment and high uncertainty in predictions.

